# Which patients did recent trials in transthyretin amyloid cardiomyopathy select? - Insights from a prospective patient registry

**DOI:** 10.1101/2025.03.11.25323803

**Authors:** Michael Poledniczek, Lena Marie Schmid, Christina Kronberger, Nikita Ermolaev, René Rettl, Christina Binder, Luciana Camuz Ligios, Mahshid Eslami, Christian Nitsche, Christian Hengstenberg, Roza Badr Eslam, Andreas Kammerlander, Johannes Kastner, Jutta Bergler-Klein, Franz Duca

## Abstract

**Introduction:** Several randomized, double-blind, placebo-controlled phase III trials (RCT) explore disease-modifying therapeutics in transthyretin amyloid cardiomyopathy (ATTR-CM). However, it is currently unclear whether patients eligible to participate in the RCT are representative of real-world patients.

**Methods:** ATTR-CM patients presenting to a tertiary referral center for cardiac amyloidosis at the Medical University of Vienna between March 2012 and May 2024 were included in a prospective cardiac amyloidosis registry. Inclusion and exclusion criteria of the ATTRACT, ATTRIBUTE, HELIOS-B, CARDIO-TTRANSFROM, and the DEPLETTR-CM trial were applied, and the baseline characteristics of the hypothetical trial cohorts as well as their survival were compared.

**Results:** A total of 353 patients (80.3 years, IQR: 75.5 – 84.2, 17.6% female) were included, out of which 192 patients would have been eligible to participate in ATTRIBUTE, ATTRACT would have recruited 163 patients, HELIOS-B 105 patients, CARDIO-TTRANSFROM 80 subjects, and 71 patients would have been eligible for DEPLETTR-CM.

Patients included in ATTRIBUTE, ATTRACT, HELIOS-B, and CARDIO-TTRANSFORM demonstrated only minor differences regarding baseline characteristics, both among each other and compared to the real-world cohort. However, patients eligible for the DEPLETTR-CM trial exhibited both more severely elevated biomarkers of heart failure (NT-proBNP: 2590pg/mL, IQR: 1614–4423, vs. 2339pg/mL, IQR: 1154–4250; p<0.001) as well more advanced disease assessed utilizing the New York Heart Association and National Amyloidosis Centre stage (p<0.001, respectively). Concerning all-cause mortality, patients who could have been included in DEPLETTR-CM also showed significantly worse survival. At the same time, no significant differences were observed between the real-world cohort and the other patient cohorts.

**Conclusions:** When applied to our real-world ATTR-CM cohort, recent RCT inclusion and exclusion criteria would have selected patients comparable to the real-world cohort. Only the DEPLETTR-CM trial would have selected patients with more advanced disease and worse prognosis in our cohort.

**Clinical perspective:** *What’s new?:*

- This is the first study to assess whether patient cohorts in recent ATTR-CM trials represent real-world ATTR-CM patients.
- While this analysis suggests that ATTRACT, ATTRIBUTE, HELIOS-B, and CARDIO-TTRANSFROM would have included a somewhat representative patient sample, DEPLETTR-CM includes patients with more advanced disease.
- Females may have been underrepresented in recent ATTR-CM trials.

*What are the clinical implications?:*

- These results may aid the interpretation of the trial results.
- This may be especially useful as currently there are no trials directly comparing ATTR-CM disease-modifying therapeutics.
- Finally, this analysis should support the design of future trials to be more representative of real- world ATTR-CM patients and more inclusive with regard to women and patients in very early disease stages.

## Introduction

Transthyretin amyloid cardiomyopathy (ATTR-CM) is an infiltrative disease of the myocardium caused by the deposition of transthyretin amyloid, resulting in cardiac hypertrophy, restrictive hemodynamics, and ultimately, heart failure [1]. Formerly considered a rare disease, the discovery of a non-invasive diagnostic algorithm [2] has increased recognition of the disease and propelled scientific and industrial efforts to develop disease-specific therapeutics.

The first approved agent for ATTR-CM was tafamidis, with results from its phase III trial published in 2018 [3]. While tafamidis has demonstrated its ability to delay disease progression by stabilizing transthyretin tetramers, disease-specific therapeutics with improved stabilization kinetics and mechanisms of action beyond the stabilization of transthyretin tetramers are emerging. These include suppressing the transcription of messenger ribonucleic acid (mRNA), inhibiting mRNA acid by hybridization, or the removal of already-deposited amyloid by induction of antibody-mediated phagocytosis [4–6].

These recently published large randomized double-blind controlled trials will give clinicians more choices for treating patients with ATTR-CM. However, no data is available to compare different disease-specific therapies against one another. With tafamidis as a well-established and safe drug, only randomized double-blinded head-to-head comparison trials would enable physicians to make an informed and truly evidence-based choice of therapy [7]. This is further complicated as the phase III trials that support the approval of agents are hardly comparable due to differences with regard to study procedures and inclusion and exclusion criteria.

We, therefore, aim to better understand patient cohorts of previously published or ongoing phase III trials by applying the respective inclusion and exclusion criteria to a real-world sample of patients with ATTR-CM. We further aim to compare the outcome of the eligible patient cohorts between trials. Ultimately, with this analysis, we seek to extend our understanding of recent large-scale phase III trials, especially whether and to what extent patients included in recent phase III ATTR-CM trials are representative of clinical characteristics and outcomes in real-world patients.

## Methods

### Setting

The present analysis was performed within the context of a prospective registry of patients diagnosed with cardiac amyloidosis at the cardiology outpatient clinic of the Vienna General Hospital, the academic referral center of the Medical University of Vienna in Vienna, Austria. The registry is approved by the local ethics committee (#1079/2023) and complies with good scientific practice guidelines and the principles outlined in the Declaration of Helsinki. All patients agreed to inclusion in the registry and provided informed consent, as evidenced by their personal signatures.

### Subjects and study design

Consecutive patients included in the cardiac amyloidosis registry between March 2012 and May 2024 were screened for eligibility and included in the present analysis if a definite diagnosis of transthyretin amyloid cardiomyopathy was established following the cardiomyopathy guidelines of the European Society of Cardiology [8].

Inclusion and exclusion criteria as defined in the first published study protocols of the 1) ATTR-ACT trial (tafamidis) [3], the 2) ATTRIBUTE-CM trial (acoramidis) [5], the 3) HELIOS-B trial (vutrisiran) [6], the 4) CARDIO – TTRANSFORM trial (eplontersen) [9], and the 5) DEPLETTR-CM trial (ALXN2220) [10] were applied. First, patients were pre-screened semi-automatically using the numerical cut-offs in the inclusion and exclusion criteria. Then, inclusion and exclusion criteria were manually applied by two cardiologists-in-training with a special research focus on cardiac amyloidosis and experience as clinical trial investigators (M.P., L.S.). First, the eligibility of all pre-selected patients was reviewed (L.S.), then these results were checked by a second investigator (M.P.). Ambiguity was resolved by consensus. In case of missing data points, e.g., six-minute walk test results at baseline, available subsequent data points, and patient documentation were analyzed. In case of documented clinical stability and/or sufficient physical capability to perform the respective tests, results were imputed by consensus of both investigators (M.P, F.D). Patients were followed for a maximum of 5 years.

### Study procedures

Demographic and clinical characteristics were compiled at the initial presentation to the institution’s outpatient clinic by a trained and board-certified cardiologist or physician in training specializing in cardiac amyloidosis. An extensive laboratory assessment, including a complete hemogram, blood chemistry, biomarkers of heart failure, electrophoresis/immunofixation of serum and urine, free light chain assay, and biomarkers of heart failure, was performed in all patients at baseline. Following the acquisition, blood analysis was performed at the central laboratory of the Medical University of Vienna, Vienna, Austria.

In addition, patients underwent a comprehensive transthoracic echocardiography examination performed by an expert echocardiographer utilizing a commercially available GE cardiovascular ultrasound system (General Electric Healthcare, Waukesha, WI, USA). Post-hoc analysis was performed using dedicated software (EchoPAC Clinical Workstation Software, Version 206, GE Vingmed Ultrasound).

### Study Endpoints

Patients were followed for 5 years from baseline, and all-cause mortality was defined as the primary endpoint. Endpoint data was compiled using 1) periodic queries on the national statistics authority’s (Statistik Austria) death registry, 2) routine out-patient visits, and 3) telephone interviews with patients or patients’ relatives.

### Statistical analysis

All categorical variables are presented as numbers and percentages, while for continuous parameters, the mean and standard deviation (SD), if normally distributed, or the median and the interquartile range (IQR) are displayed. The assumption of a normal distribution was tested using the Shapiro-Wilk test. Comparisons were drawn between all five trials, and individually between the entire cohort and the hypothetical trial cohort. Binomial or categorial parameters were compared by performing the chi-square test. For metric variables, analysis of variance or the Kruskal-Wallis was utilized as appropriate. For comparison between two groups, the t-test for independent samples or the Mann-Whitney-U test was calculated. Survival between cohorts was compared using the Kaplan-Meier method. Statistical significance was assumed with p-values of < 0.05. All statistical analyses were performed utilizing BlueSky Statistics 10.3.4, R package version 8.95 (BlueSky Statistics LLC, Chicago, IL, USA).

## Results

353 patients (80.3 years, IQR: 75.5 – 84.2, 17.6% female) were analyzed and constituted the entire study population. Those eligible for inclusion in the five hypothetical trial populations were selected out of these patients. The patient selection process is depicted in Figure 1. In our baseline ATTR-CM cohort, a total of 192 patients would have been included in ATTRIBUTE, ATTRACT would have recruited 163 patients, HELIOS-B 105 patients, CARDIO-TTRANSFROM 80 subjects, and 71 patients would have been eligible for inclusion in DEPLETTR-CM. The baseline characteristics of the entire ATTR-CM population and the hypothetical trial cohorts are displayed in Table 1 and Figure 2.

**Figure 1.**
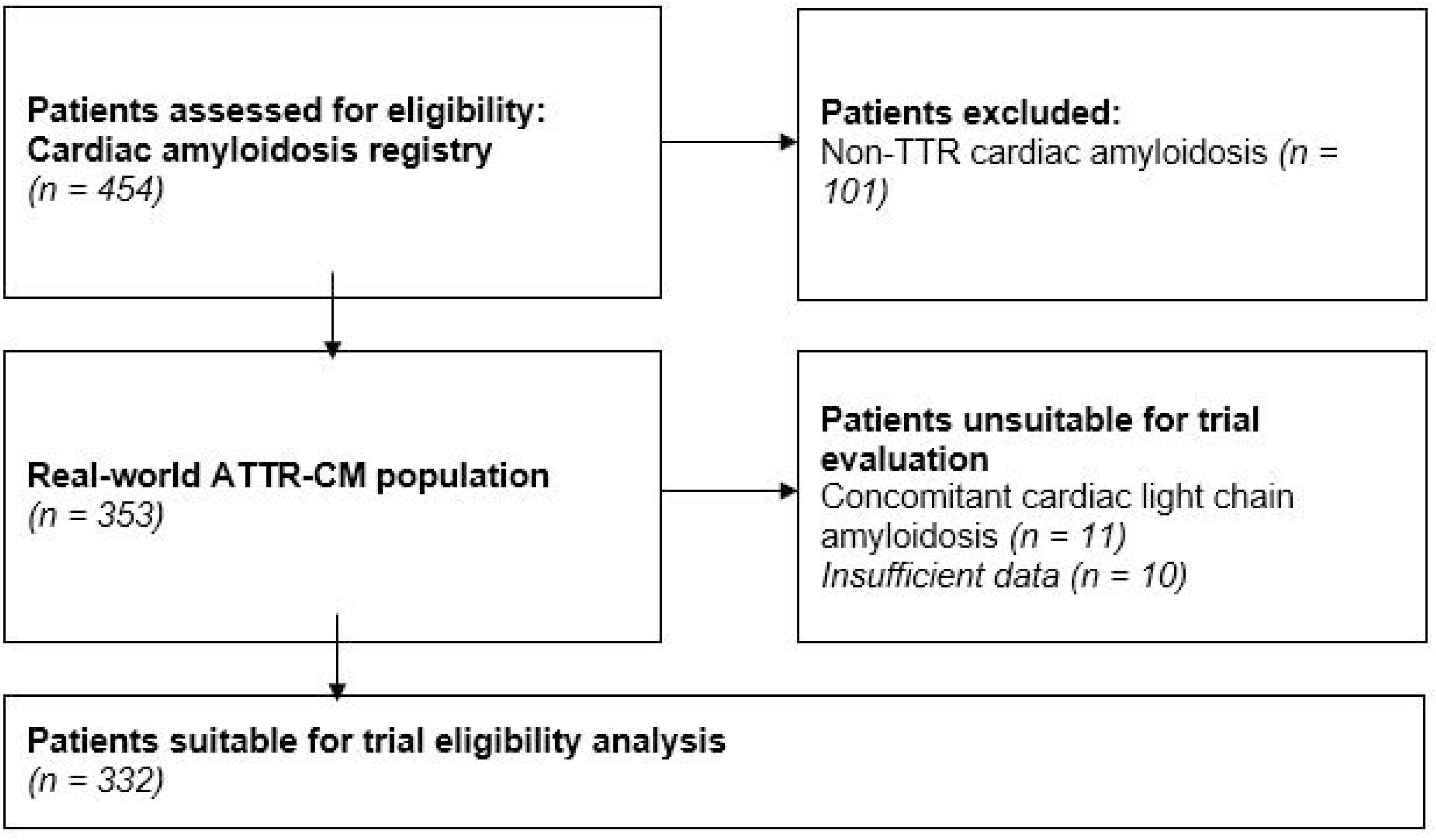
Patient recruitment flow chart. ATTR-CM, transthyretin amyloid cardiomyopathy; TTR, transthyretin.

**Figure 2.**
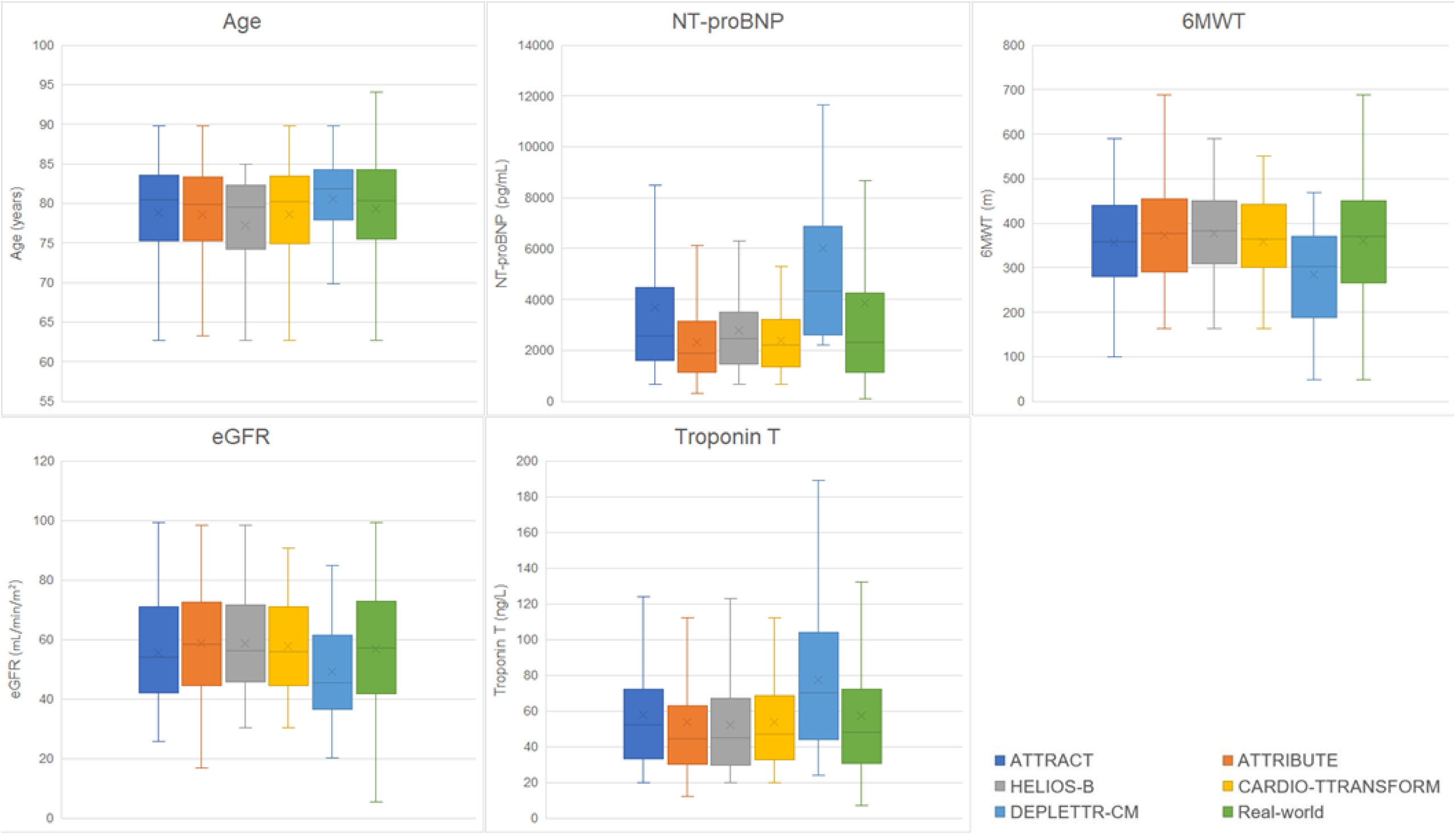
Boxplots of baseline characteristics of the entire cohort and the hypothetical trial cohorts. 6MWT, six-minute walk test; eGFR, estimated glomerular filtration rate;

**Table 1.**
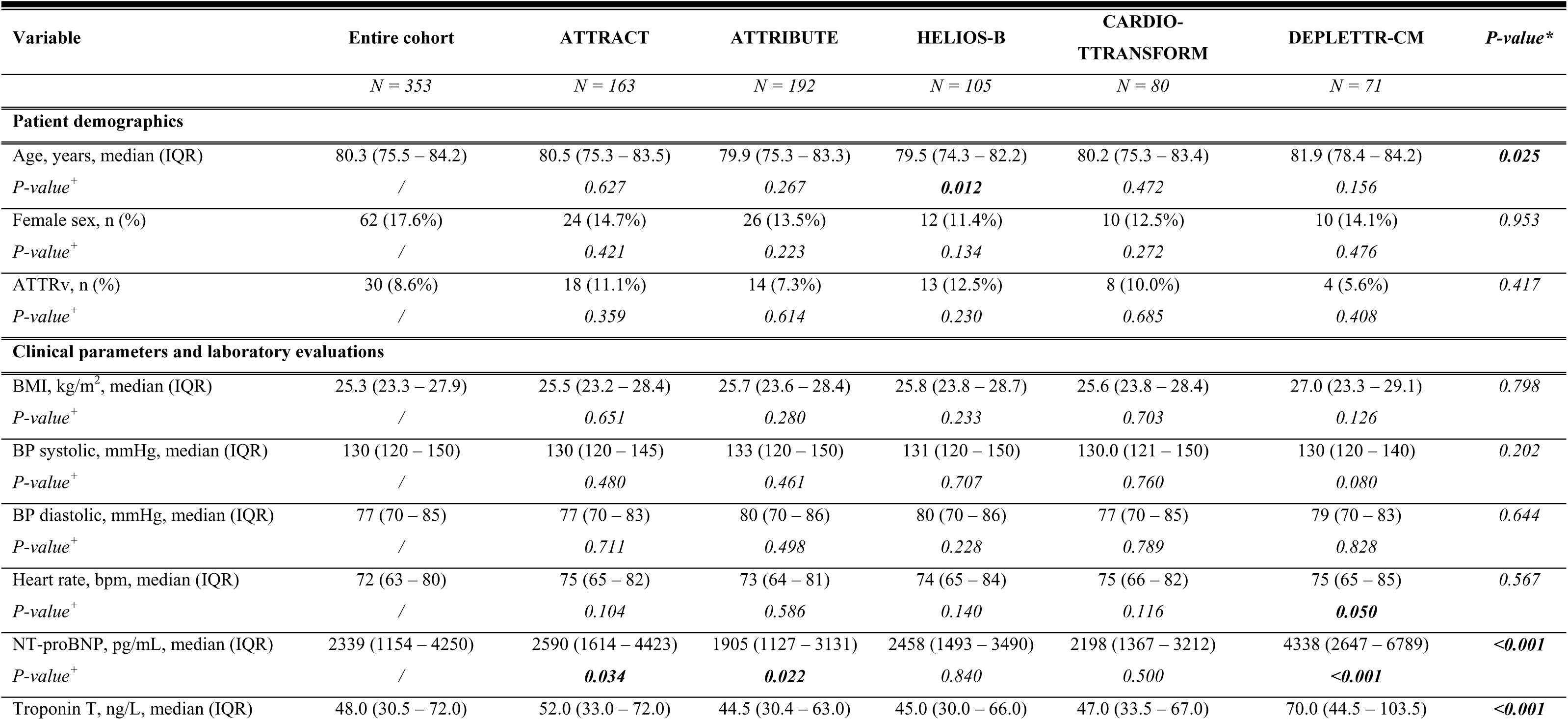
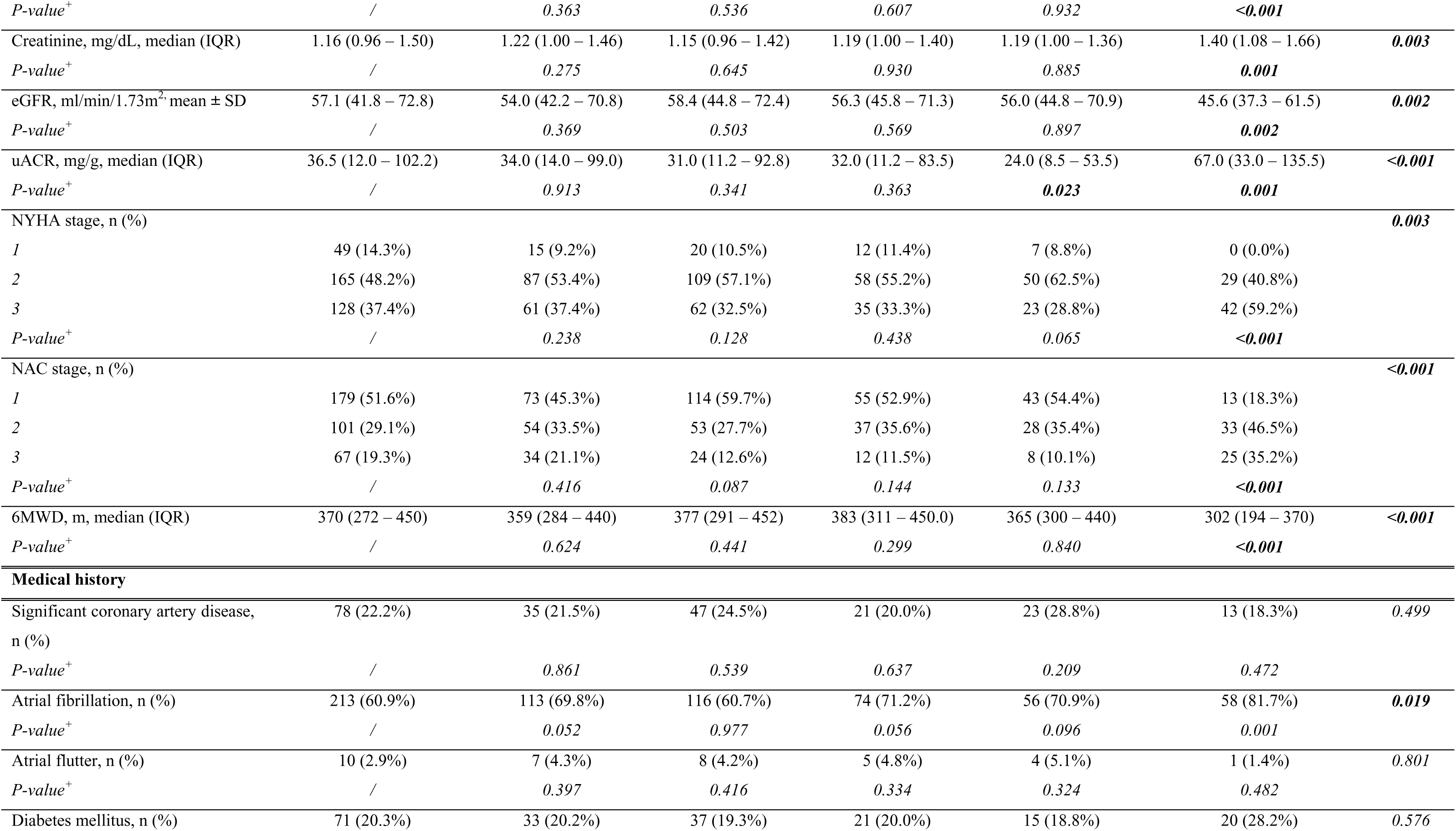
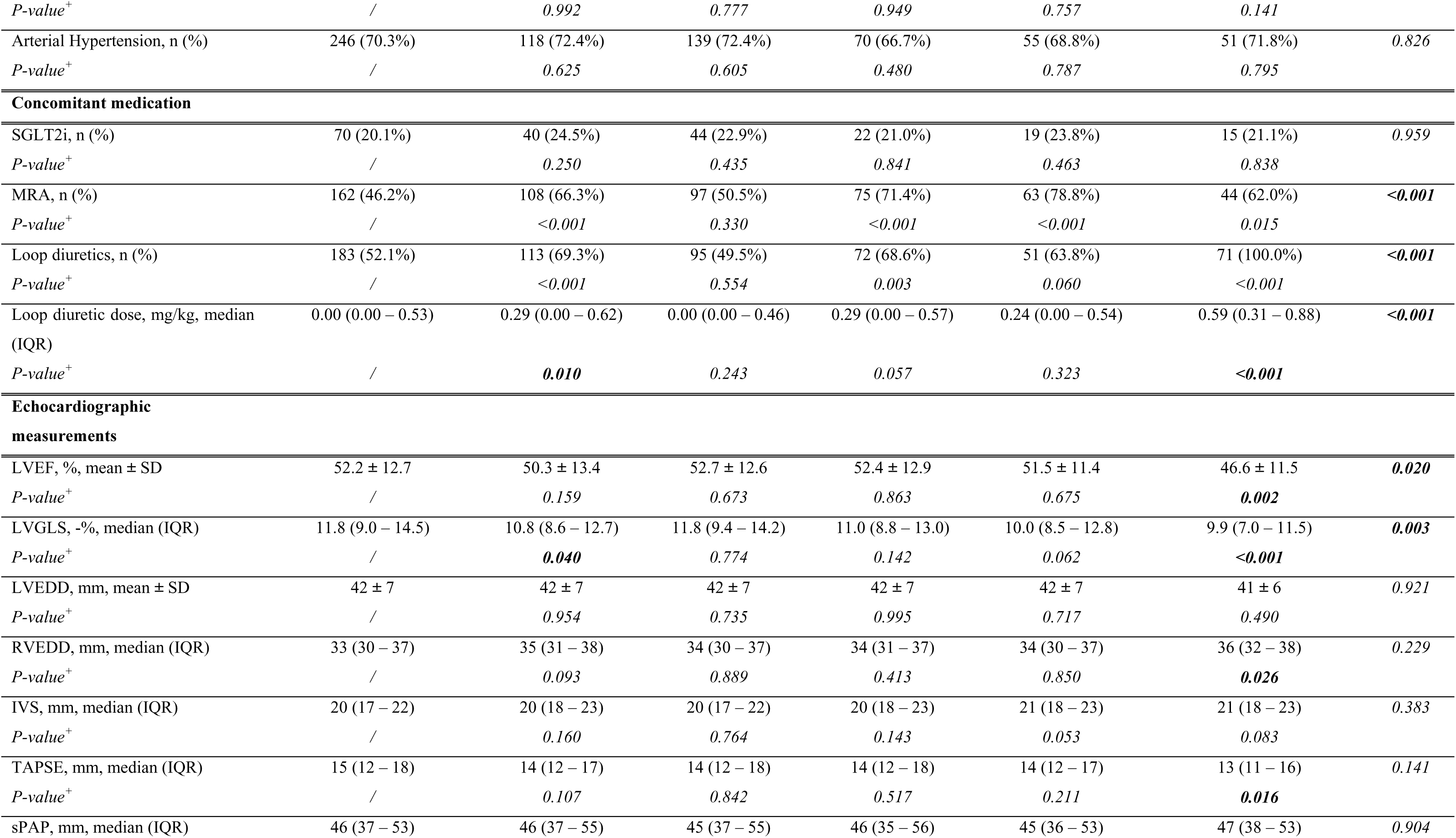
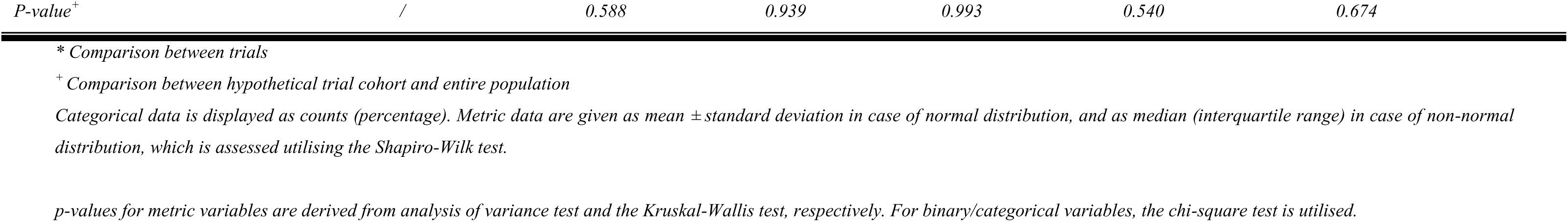
Baseline characteristics of the entire and hypothetic trial cohorts. 6MWD, six-minute walk distance; ATTRv, variant transthyretin amyloidosis; BMI, body-mass index; BP, blood pressure; bpm, beats per minute; eGFR, estimated glomerular filtration rate; IQR, interquartile range; IVS, interventricular septum; LVEDD, left ventricular end-diastolic diameter; LVEF, left ventricular ejection fraction; LVGLS, left ventricular global longitudinal strain; MRA, mineralocorticoid receptor antagonist; NAC, National Amyloidosis Centre stage; NT-proBNP, N-terminal prohormone of brain natriuretic peptide; NYHA, New York Heart Association stage; RVEDD, right ventricular end-diastolic diameter; SGLT2i, sodium-glucose cotransporter 2 inhibitor; SD, standard deviation; sPAP, systolic pulmonary artery pressure; TAPSE, tricuspid annular plane systolic excursion; uACR, urine albumin creatinine ratio;

| Variable | Entire cohort | ATTRACT | ATTRIBUTE | HELIOS-B | CARDIO-TTRANSFORM | DEPLETTR-CM | P-value* |
| --- | --- | --- | --- | --- | --- | --- | --- |
|  | N = 353 | N = 163 | N = 192 | N = 105 | N = 80 | N = 71 |  |
| **Patient demographics** |  |  |  |  |  |  |  |
| Age, years, median (IQR) | 80.3 (75.5 – 84.2) | 80.5 (75.3 – 83.5) | 79.9 (75.3 – 83.3) | 79.5 (74.3 – 82.2) | 80.2 (75.3 – 83.4) | 81.9 (78.4 – 84.2) | **0.025** |
| P-value<sup>+</sup> | / | 0.627 | 0.267 | **0.012** | 0.472 | 0.156 |  |
| Female sex, n (%) | 62 (17.6%) | 24 (14.7%) | 26 (13.5%) | 12 (11.4%) | 10 (12.5%) | 10 (14.1%) | 0.953 |
| P-value<sup>+</sup> | / | 0.421 | 0.223 | 0.134 | 0.272 | 0.476 |  |
| ATTRv, n (%) | 30 (8.6%) | 18 (11.1%) | 14 (7.3%) | 13 (12.5%) | 8 (10.0%) | 4 (5.6%) | 0.417 |
| P-value<sup>+</sup> | / | 0.359 | 0.614 | 0.230 | 0.685 | 0.408 |  |
| **Clinical parameters and laboratory evaluations** |  |  |  |  |  |  |  |
| BMI, kg/m<sup>2</sup>, median (IQR) | 25.3 (23.3 – 27.9) | 25.5 (23.2 – 28.4) | 25.7 (23.6 – 28.4) | 25.8 (23.8 – 28.7) | 25.6 (23.8 – 28.4) | 27.0 (23.3 – 29.1) | 0.798 |
| P-value<sup>+</sup> | / | 0.651 | 0.280 | 0.233 | 0.703 | 0.126 |  |
| BP systolic, mmHg, median (IQR) | 130 (120 – 150) | 130 (120 – 145) | 133 (120 – 150) | 131 (120 – 150) | 130.0 (121 – 150) | 130 (120 – 140) | 0.202 |
| P-value<sup>+</sup> | / | 0.480 | 0.461 | 0.707 | 0.760 | 0.080 |  |
| BP diastolic, mmHg, median (IQR) | 77 (70 – 85) | 77 (70 – 83) | 80 (70 – 86) | 80 (70 – 86) | 77 (70 – 85) | 79 (70 – 83) | 0.644 |
| P-value<sup>+</sup> | / | 0.711 | 0.498 | 0.228 | 0.789 | 0.828 |  |
| Heart rate, bpm, median (IQR) | 72 (63 – 80) | 75 (65 – 82) | 73 (64 – 81) | 74 (65 – 84) | 75 (66 – 82) | 75 (65 – 85) | 0.567 |
| P-value<sup>+</sup> | / | 0.104 | 0.586 | 0.140 | 0.116 | **0.050** |  |
| NT-proBNP, pg/mL, median (IQR) | 2339 (1154 – 4250) | 2590 (1614 – 4423) | 1905 (1127 – 3131) | 2458 (1493 – 3490) | 2198 (1367 – 3212) | 4338 (2647 – 6789) | **<0.001** |
| P-value<sup>+</sup> | / | **0.034** | **0.022** | 0.840 | 0.500 | **<0.001** |  |
| Troponin T, ng/L, median (IQR) | 48.0 (30.5 – 72.0) | 52.0 (33.0 – 72.0) | 44.5 (30.4 – 63.0) | 45.0 (30.0 – 66.0) | 47.0 (33.5 – 67.0) | 70.0 (44.5 – 103.5) | **<0.001** |
| *P-value*<sup>+</sup> | / | 0.363 | 0.536 | 0.607 | 0.932 | <0.001 |  |
| Creatinine, mg/dL, median (IQR) | 1.16 (0.96 – 1.50) | 1.22 (1.00 – 1.46) | 1.15 (0.96 – 1.42) | 1.19 (1.00 – 1.40) | 1.19 (1.00 – 1.36) | 1.40 (1.08 – 1.66) | **0.003** |
| *P-value*<sup>+</sup> | / | 0.275 | 0.645 | 0.930 | 0.885 | **0.001** |  |
| eGFR, ml/min/1.73m<sup>2</sup>, mean ± SD | 57.1 (41.8 – 72.8) | 54.0 (42.2 – 70.8) | 58.4 (44.8 – 72.4) | 56.3 (45.8 – 71.3) | 56.0 (44.8 – 70.9) | 45.6 (37.3 – 61.5) | **0.002** |
| *P-value*<sup>+</sup> | / | 0.369 | 0.503 | 0.569 | 0.897 | **0.002** |  |
| uACR, mg/g, median (IQR) | 36.5 (12.0 – 102.2) | 34.0 (14.0 – 99.0) | 31.0 (11.2 – 92.8) | 32.0 (11.2 – 83.5) | 24.0 (8.5 – 53.5) | 67.0 (33.0 – 135.5) | <**0.001** |
| *P-value*<sup>+</sup> | / | 0.913 | 0.341 | 0.363 | **0.023** | **0.001** |  |
| NYHA stage, n (%) |  |  |  |  |  |  | **0.003** |
| 1 | 49 (14.3%) | 15 (9.2%) | 20 (10.5%) | 12 (11.4%) | 7 (8.8%) | 0 (0.0%) |  |
| 2 | 165 (48.2%) | 87 (53.4%) | 109 (57.1%) | 58 (55.2%) | 50 (62.5%) | 29 (40.8%) |  |
| 3 | 128 (37.4%) | 61 (37.4%) | 62 (32.5%) | 35 (33.3%) | 23 (28.8%) | 42 (59.2%) |  |
| *P-value*<sup>+</sup> | / | 0.238 | 0.128 | 0.438 | 0.065 | <**0.001** |  |
| NAC stage, n (%) |  |  |  |  |  |  | <**0.001** |
| 1 | 179 (51.6%) | 73 (45.3%) | 114 (59.7%) | 55 (52.9%) | 43 (54.4%) | 13 (18.3%) |  |
| 2 | 101 (29.1%) | 54 (33.5%) | 53 (27.7%) | 37 (35.6%) | 28 (35.4%) | 33 (46.5%) |  |
| 3 | 67 (19.3%) | 34 (21.1%) | 24 (12.6%) | 12 (11.5%) | 8 (10.1%) | 25 (35.2%) |  |
| *P-value*<sup>+</sup> | / | 0.416 | 0.087 | 0.144 | 0.133 | <**0.001** |  |
| 6MWD, m, median (IQR) | 370 (272 – 450) | 359 (284 – 440) | 377 (291 – 452) | 383 (311 – 450.0) | 365 (300 – 440) | 302 (194 – 370) | <**0.001** |
| *P-value*<sup>+</sup> | / | 0.624 | 0.441 | 0.299 | 0.840 | <**0.001** |  |
| **Medical history** |  |  |  |  |  |  |  |
| Significant coronary artery disease, n (%) | 78 (22.2%) | 35 (21.5%) | 47 (24.5%) | 21 (20.0%) | 23 (28.8%) | 13 (18.3%) | 0.499 |
| *P-value*<sup>+</sup> | / | 0.861 | 0.539 | 0.637 | 0.209 | 0.472 |  |
| Atrial fibrillation, n (%) | 213 (60.9%) | 113 (69.8%) | 116 (60.7%) | 74 (71.2%) | 56 (70.9%) | 58 (81.7%) | **0.019** |
| *P-value*<sup>+</sup> | / | 0.052 | 0.977 | 0.056 | 0.096 | 0.001 |  |
| Atrial flutter, n (%) | 10 (2.9%) | 7 (4.3%) | 8 (4.2%) | 5 (4.8%) | 4 (5.1%) | 1 (1.4%) | 0.801 |
| *P-value*<sup>+</sup> | / | 0.397 | 0.416 | 0.334 | 0.324 | 0.482 |  |
| Diabetes mellitus, n (%) | 71 (20.3%) | 33 (20.2%) | 37 (19.3%) | 21 (20.0%) | 15 (18.8%) | 20 (28.2%) | 0.576 |
| *P-value*<sup>+</sup> | / | 0.992 | 0.777 | 0.949 | 0.757 | 0.141 |  |
| Arterial Hypertension, n (%) | 246 (70.3%) | 118 (72.4%) | 139 (72.4%) | 70 (66.7%) | 55 (68.8%) | 51 (71.8%) | 0.826 |
| *P-value*<sup>+</sup> | / | 0.625 | 0.605 | 0.480 | 0.787 | 0.795 |  |
| **Concomitant medication** |  |  |  |  |  |  |  |
| SGLT2i, n (%) | 70 (20.1%) | 40 (24.5%) | 44 (22.9%) | 22 (21.0%) | 19 (23.8%) | 15 (21.1%) | 0.959 |
| *P-value*<sup>+</sup> | / | 0.250 | 0.435 | 0.841 | 0.463 | 0.838 |  |
| MRA, n (%) | 162 (46.2%) | 108 (66.3%) | 97 (50.5%) | 75 (71.4%) | 63 (78.8%) | 44 (62.0%) | <0.001 |
| *P-value*<sup>+</sup> | / | <0.001 | 0.330 | <0.001 | <0.001 | 0.015 |  |
| Loop diuretics, n (%) | 183 (52.1%) | 113 (69.3%) | 95 (49.5%) | 72 (68.6%) | 51 (63.8%) | 71 (100.0%) | <0.001 |
| *P-value*<sup>+</sup> | / | <0.001 | 0.554 | 0.003 | 0.060 | <0.001 |  |
| Loop diuretic dose, mg/kg, median (IQR) | 0.00 (0.00 – 0.53) | 0.29 (0.00 – 0.62) | 0.00 (0.00 – 0.46) | 0.29 (0.00 – 0.57) | 0.24 (0.00 – 0.54) | 0.59 (0.31 – 0.88) | <0.001 |
| *P-value*<sup>+</sup> | / | 0.010 | 0.243 | 0.057 | 0.323 | <0.001 |  |
| **Echocardiographic measurements** |  |  |  |  |  |  |  |
| LVEF, %, mean ± SD | 52.2 ± 12.7 | 50.3 ± 13.4 | 52.7 ± 12.6 | 52.4 ± 12.9 | 51.5 ± 11.4 | 46.6 ± 11.5 | 0.020 |
| *P-value*<sup>+</sup> | / | 0.159 | 0.673 | 0.863 | 0.675 | 0.002 |  |
| LVGLS, -%, median (IQR) | 11.8 (9.0 – 14.5) | 10.8 (8.6 – 12.7) | 11.8 (9.4 – 14.2) | 11.0 (8.8 – 13.0) | 10.0 (8.5 – 12.8) | 9.9 (7.0 – 11.5) | 0.003 |
| *P-value*<sup>+</sup> | / | 0.040 | 0.774 | 0.142 | 0.062 | <0.001 |  |
| LVEDD, mm, mean ± SD | 42 ± 7 | 42 ± 7 | 42 ± 7 | 42 ± 7 | 42 ± 7 | 41 ± 6 | 0.921 |
| *P-value*<sup>+</sup> | / | 0.954 | 0.735 | 0.995 | 0.717 | 0.490 |  |
| RVEDD, mm, median (IQR) | 33 (30 – 37) | 35 (31 – 38) | 34 (30 – 37) | 34 (31 – 37) | 34 (30 – 37) | 36 (32 – 38) | 0.229 |
| *P-value*<sup>+</sup> | / | 0.093 | 0.889 | 0.413 | 0.850 | 0.026 |  |
| IVS, mm, median (IQR) | 20 (17 – 22) | 20 (18 – 23) | 20 (17 – 22) | 20 (18 – 23) | 21 (18 – 23) | 21 (18 – 23) | 0.383 |
| *P-value*<sup>+</sup> | / | 0.160 | 0.764 | 0.143 | 0.053 | 0.083 |  |
| TAPSE, mm, median (IQR) | 15 (12 – 18) | 14 (12 – 17) | 14 (12 – 18) | 14 (12 – 18) | 14 (12 – 17) | 13 (11 – 16) | 0.141 |
| *P-value*<sup>+</sup> | / | 0.107 | 0.842 | 0.517 | 0.211 | 0.016 |  |
| sPAP, mm, median (IQR) | 46 (37 – 53) | 46 (37 – 55) | 45 (37 – 55) | 46 (35 – 56) | 45 (36 – 53) | 47 (38 – 53) | 0.904 |

|  |  |  |  |  |  |  |
| --- | --- | --- | --- | --- | --- | --- |
| *P-value*<sup>+</sup> | / | 0.588 | 0.939 | 0.993 | 0.540 | 0.674 |

*\* Comparison between trials*
*<sup>+</sup> Comparison between hypothetical trial cohort and entire population*
*Categorical data is displayed as counts (percentage). Metric data are given as mean ± standard deviation in case of normal distribution, and as median (interquartile range) in case of non-normal distribution, which is assessed utilising the Shapiro-Wilk test.*
*p-values for metric variables are derived from analysis of variance test and the Kruskal-Wallis test, respectively. For binary/categorical variables, the chi-square test is utilised.*

In the HELIOS-B trial, patients’ median age was lower (79.5 years, IQR: 74.3 – 82.2 vs. 80.3, IQR: 75.5 – 84.2, p = 0.012). Concerning N-terminal prohormone of brain natriuretic peptide (NT-proBNP), the hypothetical ATTRIBUTE cohort exhibited lower (1905 pg/mL, IQR: 1127 – 3131, p = 0.022), and both ATTRACT (2590 pg/mL, IQR: 1614 – 4423, p = 0.034) and DEPLETTR-CM (2590 pg/mL, IQR: 1614 – 4423, p < 0.001) higher values than the real-world ATTR-CM cohort (2339 pg/mL, IQR: 1154 – 4250). The DEPLETTR-CM trial would also have included patients with a significantly higher Troponin T (70.0 ng/L, IQR: 44.5 – 103.5, vs. 48.0, IQR: 30.5 – 72.0, p < 0.001) and lower eGFR (45.6 mg/dL, IQR: 37.3 – 61.5 vs. 57.1, IQR: 41.8 – 72.8, p = 0.002) compared to the entire ATTR-CM patient population. Concerning NYHA and NAC staging, the hypothetical DEPLETTR-CM trial population would have included patients with more advanced disease (p < 0.001, respectively).

Left ventricular function assessed by left ventricular global longitudinal strain was also more severely impaired in the DEPLETTR-CM (-9.9%, IQR: 7.0 – 11.5, p < 0.001) and the ATTRACT (-10.8%, IQR: 8.6 – 12.7, p < 0.001) trials compared to the real-world sample of ATTR-CM patients (-11.8%, IQR: 9.0 – 14.5).

Figure 3 displays the Kaplan-Meier curves of the respective hypothetical trial cohorts compared to the real-world patient population. In this analysis, patients who would have been included in the DEPLETTR-CM trial had significantly worse outcomes in than the entire ATTR-CM population (p = 0.001). No significant difference in the primary endpoint of all-cause mortality was observed between the remaining hypothetical trial populations. All outcome data is depicted in Supplementary Table 1.

**Figure 3.**
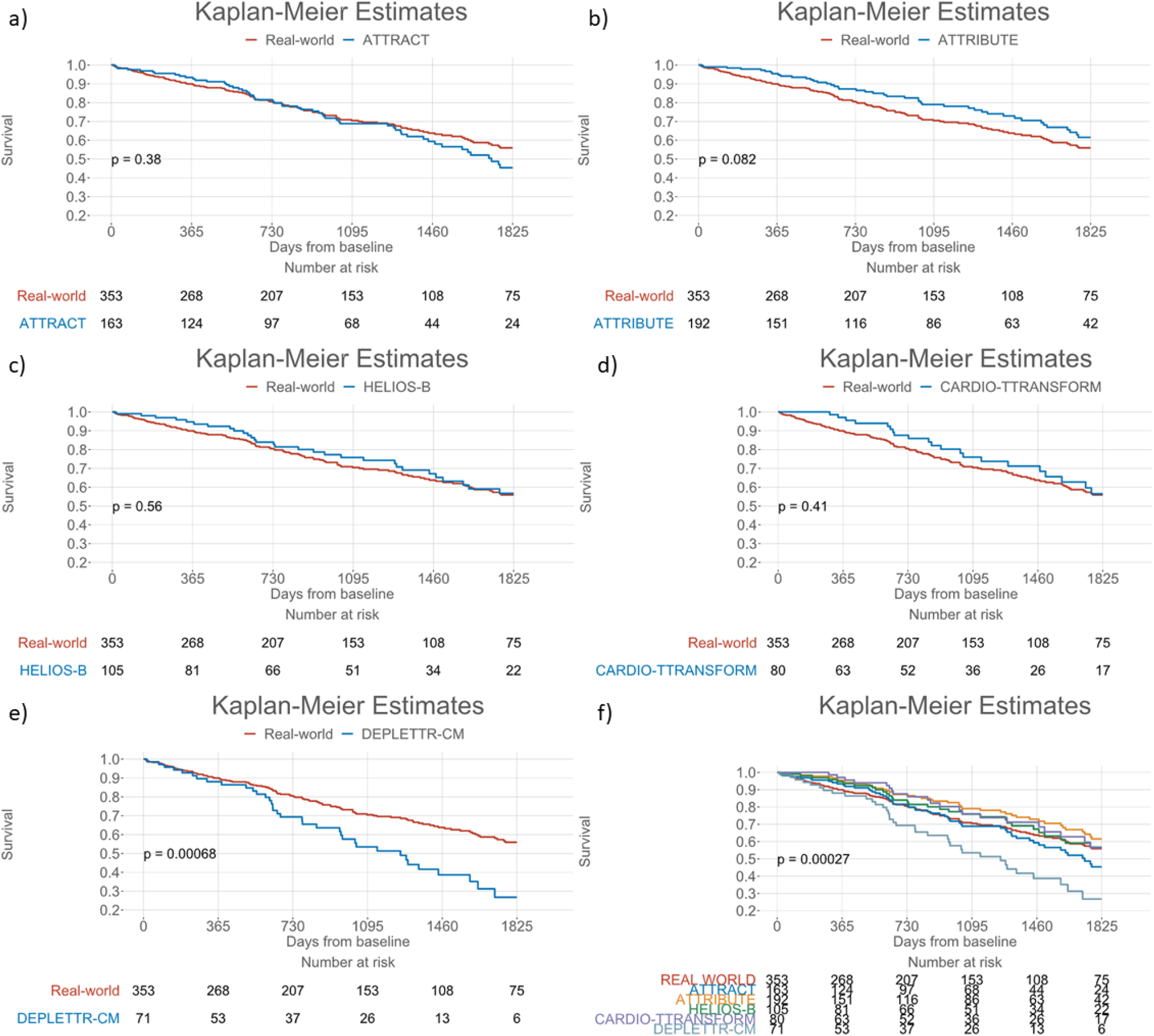
Kaplan-Meier plots of survival between a) ATTRACT, b) ATTRIBUTE, c) HELIOS-B, d) CARDIO-TTRANSFORM, e) DEPLETTR-CM and the entire real-world cohort, and f) between all trials and the real-world cohort.

Table 2 displays the published baseline characteristics of ATTRACT, ATTIBUTE, and HELIOS-B compared to patients in our hypothetical trial cohorts. In all three trials, patients recruited seem to be slightly younger. ATTRACT patients in the treatment and placebo cohort, respectively, were 74.5 (SD: 7.2) and 74.1 (SD: 6.7) years on average, while the mean age in our hypothetical ATTRACT cohort was 78.8 (SD: 7.3) years. While not statistically significant, a tendency towards lower participation of females was observed in the RCT cohorts. Furthermore, ATTRACT recruited a higher proportion of ATTRv patients [24 (24.0%) vs. 106 (14.7%), p < 0.001]. ATTRIBUTE and HELIOS-B recruited a lower proportion of NYHA stage 3 patients than would have been included in our hypothetical RCT cohort (p < 0.001, respectively). Patients included in HELIOS-B were also more likely to be in a less advanced NAC stage than our real-world ATTR-CM cohort (p = 0.002).

**Table 2.**
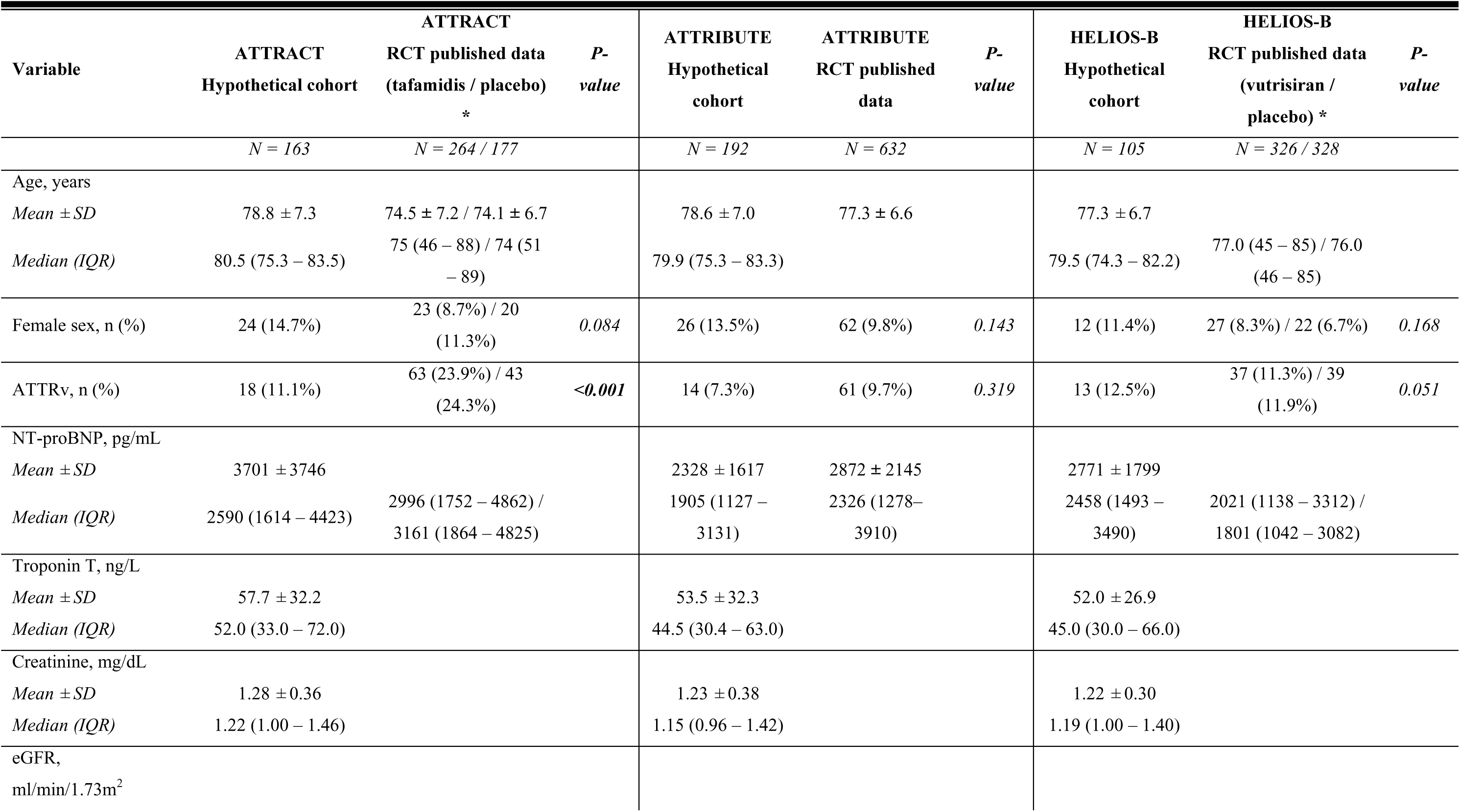
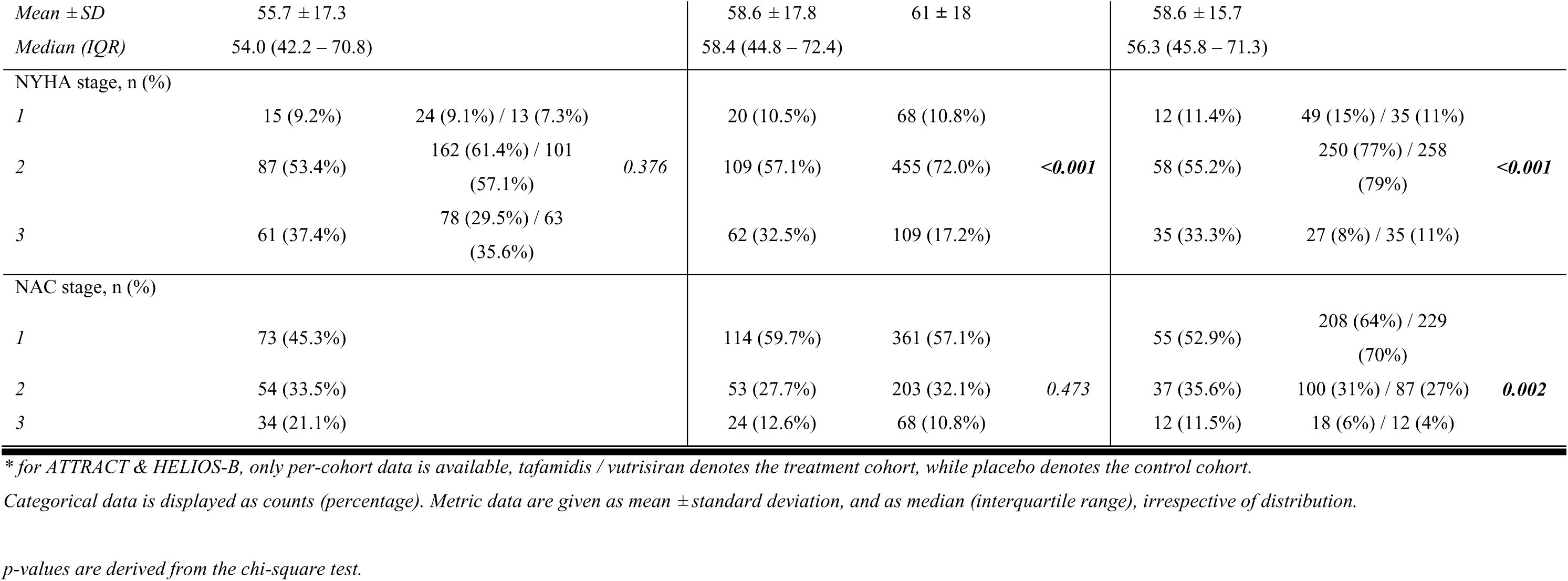
Baseline characteristics of the hypothetical and published trial cohorts. ATTRv, variant transthyretin amyloidosis; eGFR, estimated glomerular filtration rate; IQR, interquartile range; NAC, National Amyloidosis Centre stage; NT-proBNP, N-terminal prohormone of brain natriuretic peptide; NYHA, New York Heart Association stage; SD, standard deviation;

## Discussion

We have demonstrated that recent phase III trials in ATTR-CM were designed to include patient populations similar to a single-center real-world cohort. However, the DEPLETTR-CM trial seems to select patients with more severe disease.

To the best of our knowledge, this is the first study to apply the inclusion and exclusion criteria of recent RCT to a real-world population of ATTR-CM patients. Therefore, our study adds an important piece of information to aid the interpretation and understanding of recently published trial results. In addition, this analysis may also help design future trials in ATTR-CM to recruit a representative patient sample.

The most recent large-scale ATTR-CM phase III trial, the DEPLETTR-CM trial, is designed to recruit patients with the most advanced disease and worse prognosis, resulting in a more highly selected patient cohort. While this may be suspected to reduce the external validity of the DEPLETTR-CM trial results, we suggest this is a rather diligent design. As first demonstrated in the ATTRACT long-term extension, patients with advanced disease are currently not sufficiently treated with the available disease-modifying therapeutics [7], which primarily halt or slow down amyloid deposition by stabilizing transthyretin or inhibiting its production. Moreover, thus far, no treatment has demonstrated the ability to remove already-deposited amyloid from the myocardium.

On the other end of the ATTR-CM disease spectrum are patients with very early disease. Patients with NT-proBNP levels of <300 pg/mL would not have been eligible to participate in any of the trials. While DEPLETTR-CM may include patients in advanced disease stages, no trials focus on patients with very early disease. From its design, the ATTRIBUTE trial cohort is most likely to recruit such patients as the need for diuretics therapy was omitted in the exclusion criteria. Therefore, sub-group analysis of ATTRIBUTE may help to better understand the treatment effects of acoramidis as a therapeutic for patients in very early disease stages.

Results from the ATTRACT, the ATTRIBUTE, and the HELIOS-B trial have been published to date. Compared to our hypothetical RCT cohorts, these patients were less likely to be female and more likely to have a genetic ATTR variant causative of disease. While on a per-study level, this analysis failed to reach statistical significance, but for the proportion of ATTRv patients in ATTRACT, among the three trials, a highly significant tendency towards a higher proportion of male (p = 0.003) and ATTRv (p = 0.016) participants and was observed. This may raise questions about the representation of females in recent RCT and is an important issue that should be addressed in the design of future trials.

### Future research questions

Several important questions remain to be answered. First and foremost, thus far, no randomized, double-blind trials are available that compare tafamidis as the established standard of care to more recently developed gene silencers or even amyloid antibodies. Only such trials would enable truly evidence-based personalized treatment decisions and personalized ATTR-CM care.

### Strengths and Limitations

Due to the retrospective nature of our study, certain limitations need to be considered. First, with a cardiac amyloidosis registry established as early as 2014, certain changes in clinical practice occurred. The most significant change in routine diagnosis of ATTR-CM is the non-invasive diagnostic algorithm by Gillmore et al. [2], which has replaced endomyocardial biopsy in most patients. Thus, a non-invasive diagnosis was considered equivalent to an endomyocardial biopsy for the present analysis. However, a lower threshold to screen for and ultimately diagnose ATTR-CM may have altered referral patterns and baseline characteristics over time. This may be one possible explanation for the lower mean age at inclusion of the ATTRACT trial compared to our hypothetical ATTRACT cohort.

In a few patients, the six-minute walk test was not performed at baseline but at a follow-up consultation. In these cases, the stability of physical capabilities was verified based on NYHA functional class and in-hospital documentation. If sufficient evidence of clinical stability was found, these follow-up values were utilized to impute baseline values; otherwise, patients were excluded from the analysis. As this affected less than 16% of any hypothetical trial population, we are confident that this does not limit the internal validity of our findings. Furthermore, serum levels of retinol were not routinely assessed in our center. However, due to our experience with the CARDIO–TTRANSFORM trial, we think the omission of these specific exclusion criteria has not significantly changed the patient cohort characteristics for this trial.

While study participants in large phase III trials are recruited from numerous sites globally, our registry only includes patients from a single tertiary referral center, which may lead to a bias in patient selection, especially with regard to ATTRv patients. Furthermore, due to the retrospective nature of our study, certain preconditions of participation in a clinical trial could not be assessed. This includes explicitly patients’ readiness to comply with study procedures and requirements, e.g., lifestyle choices including sexual abstinence, limited alcohol consumption, and willingness to undergo intensified monitoring throughout the course of the study. It may be speculated that the preconditions to participation in such trials also introduce selection bias towards more health-educated and potentially more compliant patients. However, this is inherent to all clinical trials in cardiac amyloidosis and not a limitation to one of the included studies specifically. Rather, uncovering the differences in patient cohorts between a real-world ATTR-CM population and those included in clinical trials should drive our understanding of these randomized placebo-controlled trials and their limitations concerning patient selection.

### Conclusion

When applying our real-world ATTR-CM cohort, we can confirm that the inclusion and exclusion criteria of recent RCTs would have selected comparable patients to the real-world cohort. Only the DEPLETTR-CM trial would have selected patients with more advanced disease and worse prognosis in our cohort. Compared to our single-center real-world sample, women may be underrepresented in RCTs in ATTR-CM.

## Abbreviations

ATTR-CM: transthyretin amyloid cardiomyopathy
RCT: randomized controlled trials
NT-proBNP: N-terminal prohormone of brain natriuretic peptide
SD: standard deviation
IQR: interquartile range
NYHA: New York Heart Association functional class
NAC: National Amyloidosis Centre stage
mRNA: messenger ribonucleic acid

## Author contributions

Conceptualization: M.P., F.D.; Data curation: M.P., L.S.; Formal analysis: M.P.; Funding acquisition: n.a.; Investigation: M.P., L.S.; Methodology: M.P.; Project administration: M.P., F.D.; Resources: M.P., L.S., C.K., N.E., R.R., C.B., L.C.L., C.N.; Software: M.P.; Supervision: F.D.; Validation: M.P., L.S.; Visualization: M.P.; Writing – original draft: M.P.; Writing – review & editing: M.P., L.S., C.K., N.E., R.R., C.B., L.C.L., C.N., R.B.E., A.K., J.K., J.B.K., F.D.;

## Acknowledgments

none.

## Data availability statement

Data available upon reasonable request to the corresponding author.

## Figure legends

**Supplementary Table 1.** Outcome of the entire and hypothetic trial cohorts.

| Variable | Entire cohort | ATTRACT | ATTRIBUTE | HELIOS-B | CARDIO-TTRANSFORM | DEPLETTR-CM | *P-value* * |
| --- | --- | --- | --- | --- | --- | --- | --- |
|  | *N* = 353 | *N* = 163 | *N* = 192 | *N* = 105 | *N* = 80 | *N* = 71 |  |
| All-cause mortality, n (%) | 108 (30.6%) | 56 (34.4%) | 45 (23.4%) | 30 (28.6%) | 21 (26.2%) | 36 (50.7%) | **0.001** |
| *P-value*<sup>+</sup> | / | 0.394 | 0.076 | 0.692 | 0.443 | **0.001** |  |
| Observation period, days, median (IQR) | 939 (379 – 1651) | 943 (394 – 1496) | 973 (427 – 1736) | 1040 (434 – 1668) | 952 (528 – 1670) | 775 (368 – 1318) | 0.505 |
| *P-value*<sup>+</sup> | / | 0.688 | 0.488 | 0.536 | 0.575 | 0.179 |  |
| Time to event, days (IQR) | 650 (284 – 1036) | 776 (520 – 1298) | 812 (501 – 1319) | 736 (540 – 1290) | 862 (631 – 1292) | 661 (471 – 1066) | 0.231 |
| *P-value*<sup>+</sup> | / | 0.115 | 0.080 | 0.192 | **0.046** | 0.544 |  |

\* *Comparison between trials*
<sup>+</sup> *Comparison between hypothetical trial cohort and entire population*
*Categorical data is displayed as counts (percentage). Metric data are given as mean ± standard deviation in case of normal distribution, and as median (interquartile range) in case of non-normal distribution, which is assessed utilising the Shapiro-Wilk test.*
*p-values for metric variables are derived from analysis of variance test and the Kruskal-Wallis test, respectively. For binary/categorical variables, the chi-square test is utilised.*

